# “You just struggle on your own:” Exploring older consumers’ perspectives about falls prevention education in hospitals

**DOI:** 10.1101/2023.11.16.23298542

**Authors:** Anne-Marie Hill, Sharmila Vaz, Jacqueline Francis-Coad, Leon Flicker, Meg E. Morris, Tammy Weselman

## Abstract

**Introduction:** While individualised falls preventive education has been effective in reducing hospital falls among older patients, there is a dearth of research exploring consumers’ perspectives on hospital falls prevention education.

**Objective:** This study aimed to explore the knowledge of older consumers regarding preventing falls in hospital and their reflections on the education received during hospitalisation.

**Methods:** A qualitative, exploratory study incorporating focus groups and semi-structured interviews was undertaken. The participants consisted of a purposively selected sample of community-dwelling consumers aged 65 and above (n=39 older adults and n=9 family carers of older adults) who had been admitted to a hospital within the past five years. Thematic analysis, incorporating deductive and inductive approaches, was applied and a capability-opportunity-motivation-behaviour model was utilised to comprehend the key determinants influencing the implementation of falls education for hospitalised older adults.

**Results:** Feedback from the participants (n=46, 25 females, age range 60 to 89 years) revealed five key themes: distress and disempowerment resulting from hospital falls, anxiety and uncertainty regarding required behaviour while hospitalised, insufficient and inconsistent falls prevention education, inadequate communication, and underlying ageism attitudes. These themes converged to provide the overarching theme: “This [support and education available to engage in safe falls prevention behavior] wasn’t what we expected!” Application of the behaviour change model indicated that older consumers often did not acquire falls prevention knowledge, awareness or motivation, and had limited opportunities to engage in falls preventive behaviour during hospitalisation.

**Discussion:** In summary, older consumers receive sporadic falls prevention education during hospital admissions, which fails to raise their awareness and knowledge of falls risks or their capability to engage in safe preventive behaviours. Conflicting messages contribute to consumer confusion and anxiety about maintaining safety during hospital stays.

**Conclusion:** Further research is required to address these issues to enable older adults to undertake effective falls prevention behaviours in hospital settings. Hospital policies and guidelines need to prioritise falls prevention education. Policies can be designed to emphasise tailored education, effective communication, and the importance of addressing ageism to enhance patient safety and well-being.

## INTRODUCTION

Hospital falls are a severe, persistent, and debilitating problem to patient safety worldwide[1–3]. Up to 30% of hospital falls are associated with physical injuries [3, 4]. Typically, falls in older patients are associated with increased length of stay, functional decline, readmissions, and considerable costs to individuals, hospitals, healthcare providers, and insurers [5–9].

Health services use falls prevention interventions that show limited evidence of effectiveness [10]. A recent systematic review found that commonly used hospital falls prevention interventions such as chair alarms or wearable sensors showed little effect in reducing falls [11]. The review found that multifactorial interventions that included combinations of at least two interventions of patient or staff education, procedures for nurse handover, fast responses to patient call bells, regular toileting, environmental modifications, assistive devices, exercise therapies, safe footwear, medication management, diet or management of cognitive impairment showed some promising effect. Educating patients and staff about the risk of falls and suitable strategies for mitigating falls was the only intervention that significantly reduced fall rates and older peoples’ risk of falling in hospitals [11]. The recent World Falls guidelines have now recommended educating patients to mitigate fall risk (Grade 1A evidence) [12].

In routine practice, staff deliver fall prevention education to hospitalised patients [13]. Patient falls prevention education has evidence of effectiveness when delivered as a ward-level intervention with staff support [4], and evidence suggests that directly educating patients assists in positive safety cultures in hospital wards [14–16]. However, scant, systematic implementation of patient falls prevention education has been reported [17]. Despite patients being at highest risk of sustaining a fall in the first week of their hospital admission [18, 19], it remains a challenge for health professionals to adjust their practice to prioritise delivering fall education to patients soon after admission [20, 21].

Despite sound theoretical basis and empirical support, many interventions do not produce real-world change because few are successfully implemented [22]. The impact of failed implementation in hospitals could be far-reaching, including a reduction in the quality of patient care and reluctance of staff to try alternative interventions if they had an unsuccessful experience [23, 24]. Social marketing concepts in health show that a consumer focus is essential to motivate behaviour change [25]. Before implementing any consumer-focused education program, it is important to explore consumer experiences. For example, engaging older adults in fall prevention education is believed to empower them to take responsibility for being safe [26]. Consulting with consumer groups and aligning interventions with their views aids in tailoring future programs and validates consumers’ lived experiences and insights, thus making them more willing to engage with and adhere to the intervention [27].

This paper reports the first phase of a project that aimed to understand how to effectively implement patient-focused falls prevention education in hospitals. In this first phase, feedback from older consumers was sought to understand what they already knew about hospital falls prevention and their current experiences with inpatient falls prevention education.. The objective of the current study was to explore older consumers’ knowledge about hospital falls prevention and their reflections about falls prevention education they received when hospitalised.

## METHODS

### Ethical considerations

Ethics approval for the study was obtained from the University of Western Australia, Human Research Ethics Committee (2022/ET000692). All participants provided written informed consent to participate.

### Design

Exploratory, descriptive qualitative approaches entailing focus groups and semi-structured interviews were used to address the aim [28–30]. Focus groups were used to foster spontaneous and honest responses and help unpack pragmatic strategies to address the aim [31, 32]. Semi-structured interviews helped obtain an in-depth exploration of older adults’ reflections and feedback, validate the focus groups’ findings, unpack new themes, and triangulate findings through member checking [33].

The study was guided by the Capability-Opportunity-Motivation-Behaviour model (COM-B) [34, 35]. The COM-B model is widely used to identify what needs to change for a behaviour change intervention to be effective. COM-B has been depicted in several frameworks, varying in the specification of the causal links between capability, opportunity, motivation, and behaviour [34, 35]. The differences arise from the different levels of elaboration of the model. At its simplest level, COM-B postulates that for a person to engage in health behaviour (in this instance, falls preventive strategies in a hospital setting), three factors need to be present: **C**apability (both physical and psychological, including knowledge and awareness about the problem of falls and suitable preventive strategies), **O**pportunities afforded by the social and environmental conditions (such as being provided with relevant equipment and staff support to engage in the relevant behaviours), and **M**otivation (reflective OR automatic) to undertake the behaviour (engaging in personally relevant falls preventive strategies). COM-B developers have suggested that there can also be causal influences from Motivation to Capability and Opportunity and for greater precision, the contribution of Capability and Opportunity on the relationship between Motivation and Behaviour rather than behaviour directly should be explored [34]. Adding an interpretive dimension to the study by framing it around the COM-B model, the researchers aimed to inform, support, and challenge the current education practices for falls prevention in hospitals.

### Setting

The study was conducted in Western Australia (WA), which provides hospital care through various settings ranging from large tertiary hospitals to regional and small rural settings. As part of system-wide quality and safety services, all WA hospitals provide structured and systematic falls prevention programs that inform hospital care for all admitted patients. Older people may also develop knowledge and awareness about falls prevention through programs that operate locally and nationally in the broader community.

### Participants

Inclusion criteria were having a history of being admitted to a hospital, having experiences related to falls, including falls in hospitals, being able to provide informed consent, communicating in English and being 65 years or older. Caregivers of older adults were eligible if they had carer-related experiences related to the older adult they cared for being admitted to a hospital.

### Recruitment and sampling

Purposeful sampling was used to recruit community-dwelling older adults and their caregivers. The sampling strategy sought to maximise variation in participant characteristics of age, gender, type of hospital experience, and cultural background and health status, reflected by characteristics such as the number of medications taken (indicating multimorbidity, while four or more medications indicates increased risk of falls [36] and use of a walking aid (indicates reduced functional ability and increased risk of falls [37]. The research team liaised with a local not-for-profit WA organisation to assist with promotion and advertising [38]. Potential participants responded to the advertisement by phoning or emailing the research team, who presented written information about the study and obtained informed consent. No eligible consumers who responded refused to participate.

### Data Collection

Participants were given the choice to attend a focus group or a one-on-one interview either in person at a designated quiet, private location within a community centre or online using Microsoft Teams application (Version 1.6.00.11166). Participation options were provided because some older adults living with chronic diseases, expressed reservations about attending groups due to possible infections like COVID-19. Focus groups lasted two to three hours and included three to ten participants. Two to three researchers (AMH, TW, SV) facilitated each focus group with shared moderation and observation, with the observer taking notes. One researcher (TW), with support from AMH, conducted individual semi-structured interviews, which took 45 to 60 minutes. At the conclusion of interview and focus groups, the researchers summarised the results from their perspective and asked participants if it reflected their understanding of the feedback. Participant demographic data was collected via a short survey completed at the beginning of the interview or focus group. The researchers were experienced health professionals, all had research training, and two had educational qualifications. All researchers had worked directly with older people in research projects for at least 15 years, and one of the researchers was an older adult. The researchers conferred throughout and interviews ceased when they agreed that data saturation had been reached, with no new data being generated and data gathered fitting into previously described themes [28].

### Interview topic guide

Interviews and focus groups used a discussion guide that was followed to facilitate and steer the conversation. The authors used the COM-B model to explore older consumers’ knowledge about hospital falls prevention (using an inductive approach) and understand (deductively) if they were aware of falls prior to or when hospitalised and whether they were given suitable information and motivated to engage in safe behaviours or received support when hospitalised; and were given opportunities to engage in falls risk reduction behaviours.

### Data analysis

Demographic data were summarised using descriptive statistics. All qualitative data were audio recorded, transcribed verbatim and managed using NVivo (QSR International Pty Ltd.). Transcripts were manually checked by two researchers (TW, AMH) for accuracy and completeness [29]. Thematic analysis using an inductive approach generated themes regarding people’s falls knowledge. A six-phase guide to reflexive thematic analysis was followed [39, 40]: familiarity with the data set; generating initial codes; searching for themes; reviewing candidate themes (and producing a ‘thematic map’); defining and naming themes, and producing the analytical report presented here. A deductive approach was used to code responses by theme and assigned to the pre-determined COM-B model themes of capability, opportunity, and motivation. Subcategories with similarities were grouped within each generic category and then grouped under higher-order headings to reduce the number of categories through the collapse of like and unlike categories. Analysis began during the data collection phase to support decision-making regarding sampling and saturation [28].

Several steps were taken to enhance the rigour and transparency of data collection and analyses, as well as documenting the active engagement of the researchers with the data – in line with the reflexive thematic analysis approach [40]. Two researchers (TW and AMH) independently coded an initial sample of transcripts, and subsequently collaboratively identified candidate themes, with ongoing discussions of coding and themes with a third senior researcher (JFC) throughout the analysis. This process ensured a collaborative, nuanced analysis of the data by the research team – as opposed to merely seeking a shared consensus on meaning [40]. An audit trail was followed to enhance dependability, enabling the researchers to identify links between raw data and final findings and decisions made throughout these inductive and deductive processes [28].

## RESULTS

### Participant characteristics

Twenty-two older consumers and eight carers in two areas of the Perth metropolitan region participated in three focus groups, and a further 16 participants, (including one carer), completed an individual semi-structured interview (*N*=46 participants). The characteristics of the sample are presented in Tables 1 and 2. Briefly, of the 37 older consumers 26 (70.3%) had been admitted to a large tertiary hospital, and 21 (54.1%) had more than one hospital admission. Seven participants (18.4%) reported they fell or almost fell while hospitalised, and 25 (65.8%) participants had a fall at home in the previous year.

**Table 1.**
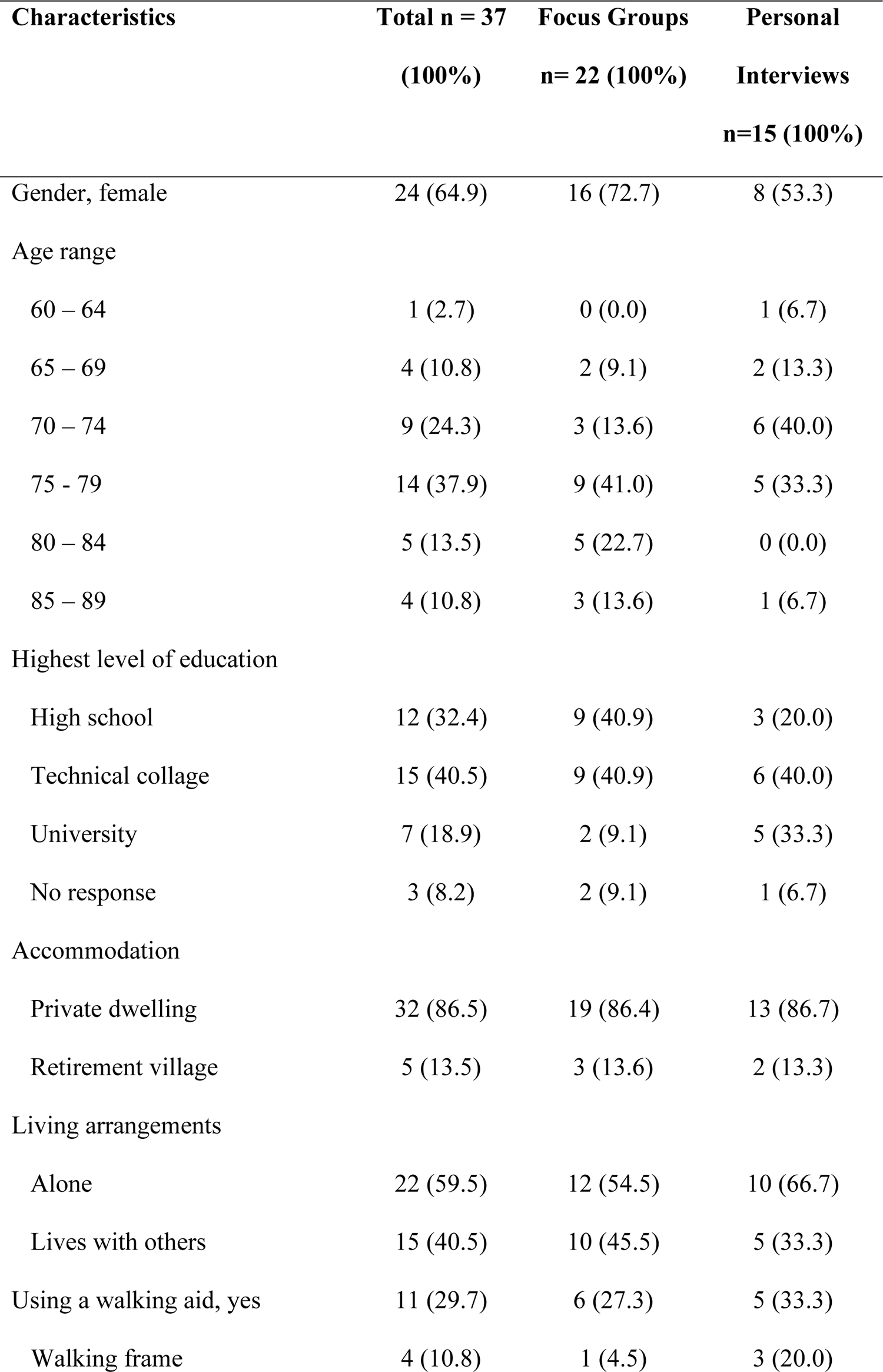
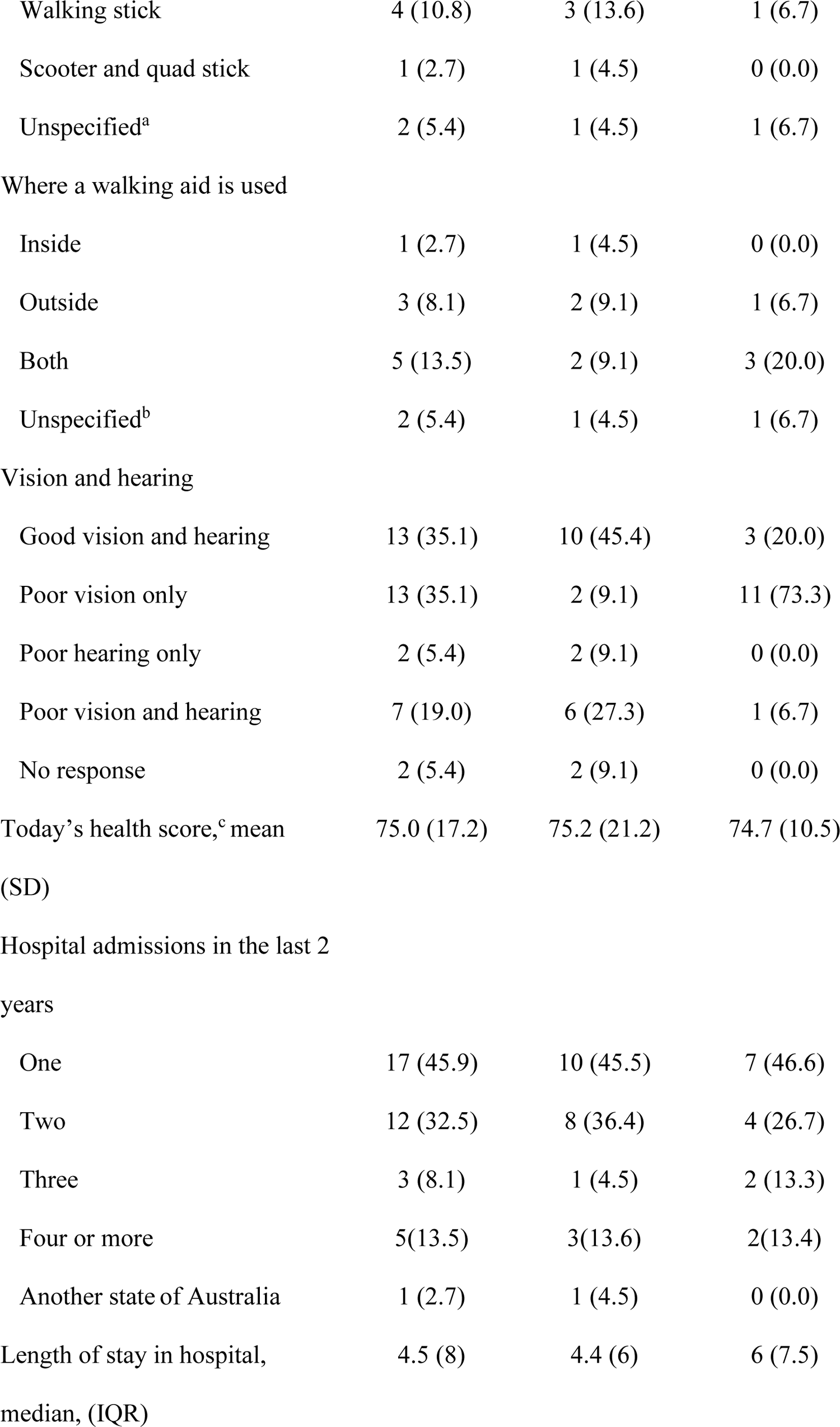
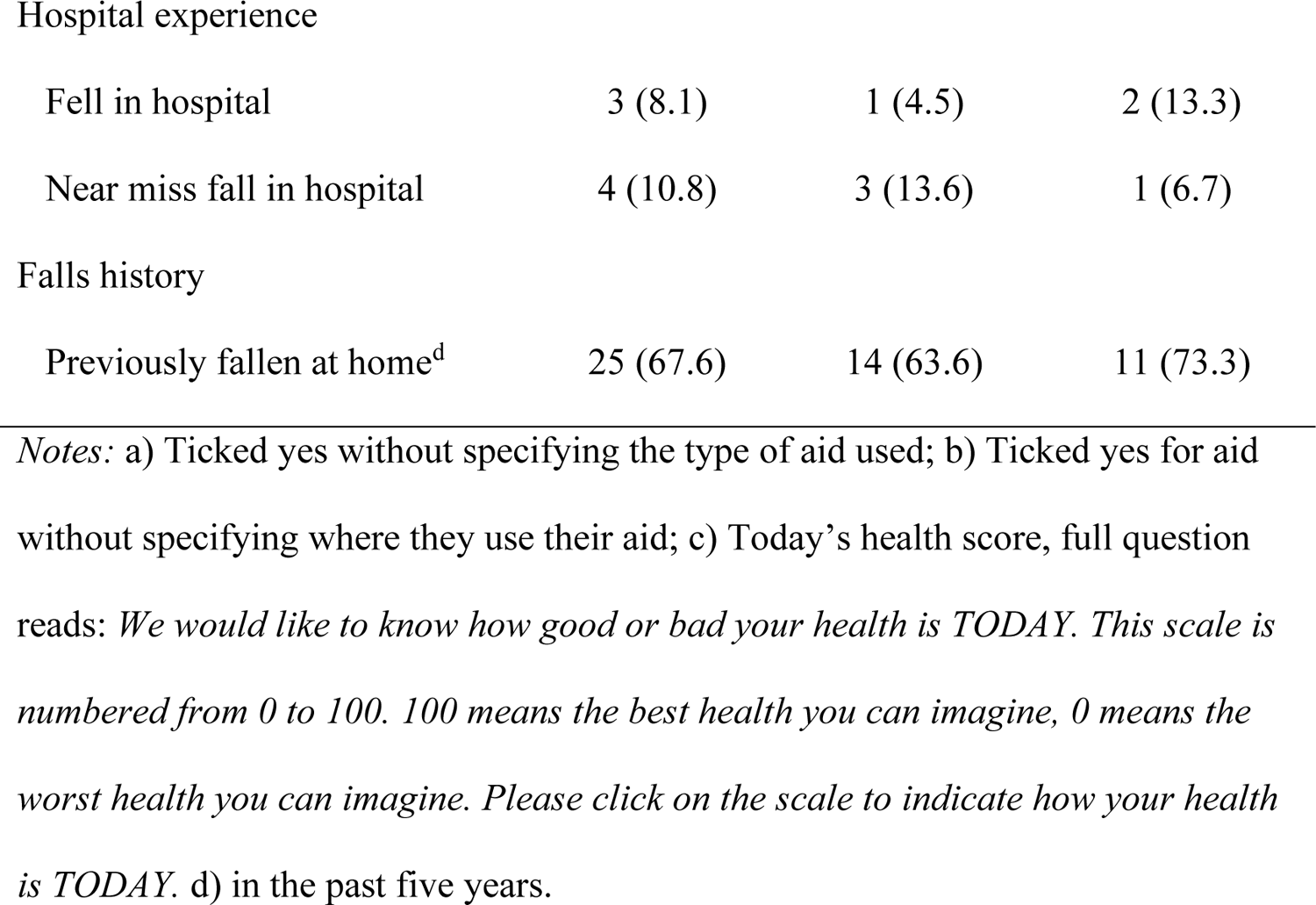
Older adult sample.

**Table 2.** Carer sample.

| **Characteristics** | **n = 9<sup>a</sup>** |
| --- | --- |
| Age, years, range |  |
| less than 54 | 4 |
| 55 - 59 | 2 |
| 60 - 64 | 1 |
| 65 - 69 | 1 |
| 75 - 79 | 1 |
| Gender, female | 8 (88.8%) |
| Person caring for: |  |
| Daughter caring for mother | 4 |
| Daughter caring for father | 3 |
| Husband caring for wife | 1 |
| Caring for more than one person<sup>b</sup> | 1 |

*Notes:* a) Eight carers attended focus groups, one was interviewed; b An Aboriginal person who cared for multiple people in her community

### Consumer’s reflections regarding falls education in hospitals

Insights from consumers identified five themes that reflected their feedback about the falls prevention education they encountered in hospitals: (1) the distress and disempowerment experienced from hospital falls or near-falls, (2) the anxiety and uncertainty surrounding expectations about engaging in falls preventive behaviour in hospital, (3) insufficient and inconsistent falls prevention education provided, (4) inadequate communication and (5) underlying attitudes of ageism. These themes converged to provide the overarching theme, “This [support and education available to engage in safe falls prevention behaviour] wasn’t what we expected!” The overarching theme reflected the emotional vulnerability of older consumers and their carers due to falls, their uncertainty about what was expected and the lack of opportunity to initiate safe behaviours while in hospital.

### Theme 1. Distress and disempowerment caused by falls

#### Sub-theme: Falls are a distressing experience

Participants personal or family experiences of falls in the hospital engendered feelings of physical and psychological distress. Older consumers vividly recalled the heightened trauma of the fall. A daughter described her mother’s experience by stating “…her last fall was really major, in the bathroom…she cracked her skull…she did some pretty serious damage” (P1, carer). Another participant recounted her anguish from a fall she experienced whilst awaiting surgery, stating, “…I was told that we’ll [*staff on the ward*] give you a couple of really strong pain pills…that was fine, and I went into bed. Then I got up from bed again to go to the toilet…when I was coming back, I had a fall…near my bed…and hit my forehead on the ground” (P11).

#### Sub-theme. Falls can result in disempowerment and helplessness

Many consumers described experiences of disempowerment and limited autonomy after a fall or when trying to prevent themselves from falling during their hospital stay. One participant described having his movements restricted after sustaining a fall in the hospital, “I had a fall trying to get outta bed, uh, on, on the first night… uh, and they used a, a hoist to get me up off the floor and then put an alert mat on the bed…so I wouldn’t get out of bed again” (P14). A carer (daughter-in-law) recounted feelings of helplessness as her family members wanted to assist their mother who was at high risk of falls, “…she did have a few, you know where she did fall … or she didn’t fall but she got very wobbly on her feet and there was never really anybody there to help her (P1).” A family member, whose father incurred a serious head injury from a hospital fall, recounted feeling powerless and angry regarding her post-fall experience, “…so I was ringing half an hour, every half an hour to find out what was going on, and I said to the reception, I said, if I don’t speak to someone now I’m putting a complaint in right now” (P40, carer). Another participant described feeling disempowered and uncared for while trying to mobilise safely.

“I needed to get out to the toilet…and I could not feel a bell anywhere. And when I said that next morning, they said, oh, it just fell off. It was on the floor…to make things worse, I could hear a few [staff]…. laughing and talking and carrying on … and nobody even found the bell on the floor for me” (P11).

### Theme 2. Uncertainty about how to behave safely while in hospital

Unfamiliar hospital environments contributed to patients’ feelings of heightened fall risk and reduced opportunity to engage in fall prevention strategies. Patients expressed feelings of anxiety and uncertainty about what they should be doing and how to behave.

#### Sub-theme. Unfamiliar, busy hospital environments and risk of falls

Participants described how the opportunity to engage in safe behaviour was compromised due to unfamiliarity with the environment and personal context. One participant recalled his father falling in hospital: “…he got up in the middle of the night to go to the toilet, and he had a catheter in, he didn’t think, obviously, about stuff that was going on” (P28). Multiple consumers perceived that ward environments were consistently busy and hence not conducive to personalised assistance even when patients thought it was required. “From my experience, when they’re doing handover, seven o’clock in the morning usually if you needed help, there was no point ringing the buzzer ‘cause they weren’t gonna come” (P14). Staff behaviour on the ward was also perceived to impact directly on patient safety, as one carer stated that “…some nurses made sure the call bell was right there, but not everyone did. No, (for) my dad, every time I went in, the first thing I did was scanning… right, where is the bell? Where is it? And sometimes he couldn’t reach it” (P39, carer).

#### Sub-theme: Medical problems impacted negatively on capability

Consumers perceived they were not always physically or psychologically capable of safely engaging in their care while in hospital. “I was off my game at the beginning after being admitted” (P16). Another participant stated, “…I had no idea what day of the week, much less the month or date” (P32). One participant described the problem of being dizzy when unwell, stating that “…one of the things that does happen is people have reached for their walking aid and slipped over. Yeah, too dizzy” (P34).

#### Sub-theme. Feeling deterred from seeking assistance

Motivation cues were described vividly by most older consumers as deterring them from seeking assistance. Participants’ strong perceptions were of staff being ‘too busy to help’ and some patients expressed feelings that their needs were of low priority for staff attention. One participant reflected that “…you press the bell, you wait your turn, because the nurses have got other jobs to do, you gotta wait” (P30) and another stated, “…I mean they do their best, but having been in there… (I) know how many times that bell gets ignored” (P13).

Older consumers consistently identified an apparent paradox between needing help and seeking help, confusing them about how to remain safe on the ward, with one stating: “I know that people are hesitant to call a nurse in hospital because nurses are busy” (P33). Some participants were aware that lack of staff assistance was compromising their safety. “I was told if I wanted to go for a walk to ask for it, but the staff is so short that they’re helping me walk around when somebody else really needs treatment, so you don’t do it and you just struggle on your own” (P13).

Overall, consumers expressed an understanding that staff tasks might lead to limitations in interactions and timely assistance. One consumer stated “…I appreciate the nurse’s job is they don’t have a moment to spare…they work very hard” (P5), while another commented, “…when they’re busy, they don’t often get to the things that are lower down on the list. You know, they have to make priorities” (P38, carer). Alternatively, some consumers perceived that staff were sometimes angry and disrespectful. One participant began to cry as she described her experiences of communication with staff.

> [Crying while talking] “I did have an infected knee which was so painful…unbearable… and a nurse brought me the commode and said, it’s there for your comfort if you need. And, then another nurse came… I believe she was the head nurse. And she said, who put that there? And I said, well, one of your nurses did. Then she says, you don’t use that here. This nurse spoke to me like this [participant speaking in a stern voice to show the group how the nurse spoke to her]. you stand, and you use the toilet that’s in the bathroom. And I was in so much pain… I rang my husband up in tears…it was shocking.(P29)”

#### Sub-theme. Feeling compelled to engage in unsafe behaviour

Untimely responses to ringing the call bell for assistance, particularly for going to the toilet, was viewed as being forced to take action (no choice) alone, even if the consequences were detrimental to safety, creating a ‘detriment versus dignity’ situation. Participants emotionally described undignified toileting experiences. One participant stated “…I don’t know, but having a pee or poo in your pants isn’t a great feeling. Absolutely not! It’s so degrading” (P14). Another explained her frustration: “It doesn’t make sense to me, if you ring the bell for the toilet and they don’t show up for half an hour…What do you do? completely lose your dignity?” (P11).

### Theme 3. Inadequate education to develop capability and motivation

Most participants did not recall any information being provided that would have developed their capability (knowledge and awareness) to engage in falls prevention behaviours while in hospital.

Sub-theme. Surprised to hear that falls prevention is important when in hospital Participants were consistently surprised to be asked about their recall of receiving hospital falls education. Multiple participants in at one focus group table stated emphatically (with nodding from all participants) “No” (P2) or “Not me” (P6). One interviewee stated “I don’t recall anything to do with falls at all” (P28). Another participant reflected “…I spent all this time in hospital…I didn’t see one thing about falls. Nobody even spoke to me about falling, so I wasn’t aware that it was an issue” (P13). One consumer summarised their falls education experience over multiple hospital admissions by stating, “…I don’t think it [how to prevent falls in hospital] was explained very well. And I was back and forth to the hospital for about a year” (P20).

#### Sub-theme. Sporadic information or education about falls prevention

A few participants recalled specific information about falls being provided with variation in the content and quality. Participants who received verbal falls prevention information from a health professional described it as a highly positive experience that had assisted their safe behaviour. One consumer recounted what the physiotherapist had said, “…I want to explain to you what you can and can’t do right from the beginning. The hospital physio was very good…very clear” (P20). Another participant recalled being given clear instructions about ringing the bell for assistance, stating “…[staff] did say if I felt I wasn’t able to do it [get up safely] to buzz them and they would come and assist” (P31).

Participants’ experiences of educational falls prevention resources in hospital, such as signs, charts or brochures to assist them being safe were mixed, with one stating “…the information [provided] in hospitals is a bit overwhelming” (P34), while another participant stated “…there was an information book at the bedside” (P16).

#### Sub-theme. Need to raise consumers’ awareness about falls in hospital

Participants spontaneously extended their reflections to describe the dangers of older patients’ heightened risk of falls while in hospital and were cognisant of the need for more direct awareness raising.

> “The self-realisation or whatever you need to do… I just think that would not be occurring for people…they just would assume they felt OK before they came into hospital. They’ve felt OK lying in bed and…they don’t factor in the medication or the sedation or anaesthetic or whatever” (P34).

This reflection was supported by other participants who commented, “…sometimes after, well a stroke or maybe even after surgery or some illness.. patients often overestimate their ability to move…which is dangerous” (P14). One consumer expressed that there was a mismatch between staff and older patients about the need for help with more overt communication required. “I sort of think expecting people to know they need help is the biggest problem… older patients have to be told you cannot get out of bed without help for the first 24 hours or something” (P34).

#### Sub-theme. Prior learning as an older adult

Participants’ knowledge and decision-making about reducing their risk of falls in hospital was predominantly based on their prior learning from experiences about falls as an older adult in the community. Nearly all participants expressed some knowledge that contributed to their current capability to engage in falls prevention. A participant commented that he had learned how important it was to be careful at night, “Because it’s a dark room you’ll be more conscious of the fact.. you could hurt yourself and be more careful” (P5). Another recounted her safe actions at home: “I can’t rush as much as I used to…to dress, I have to sit and then put on my clothes” (P19). Other participants were cognisant of falls being a problem associated with ageing. “I’m very aware that I’m not as stable as I’m used to be and so I’m probably overcautious…” (P31).)”. One participant expressed a view, that as an adult learner, older patients should take personal responsibility for falls prevention “I couldn’t help thinking it was just a bit one-sided… fall prevention is always the patient’s responsibility and I know that the staff wouldn’t think that, but it’s gotta start with yourself.”(P32).

### Theme 4. Communication is inadequate

#### Sub-theme. Inconsistent and confusing communication

Multiple participants spontaneously expanded their reflections about receiving inconsistent falls prevention education and their confusion about how to behave while in hospital to suggest that an underlying problem in hospitals was inadequate communication. Feedback included not being listened to and not receiving relevant and important information. One patient described her feelings by stating “you press the button, but you have no way of letting anyone know that you’re desperate…” (P23), while another stated that the main problem was “…not being given any information, not being told what was going on…” (P11). One participant commented that “…when I went into the ward, the information was important, but was behind the patient’s head on the wall so the patient couldn’t see it!” (P34). Another patient expressed frustration at her knowledge being ignored, stating “…nobody would listen to me because they didn’t have the paperwork – it was not sent from another hospital so they wouldn’t listen…” (P3).

#### Sub-theme. Essential safety information not presented

Inadequate communication included relevant safety information not being explained to the patient. An older consumer recounted how there was a bedside communication board to promote safe assistance, but it was not useful because staff did not keep it updated. “…it was in the ward…and the nursing staff is supposed to write on it - who’s looking after you and comments – but it never got used …. nobody paid any attention to it” (P13). Another consumer carer observed that her father could not benefit from the bedside communication board because he was unaware that it recorded important mobility actions that could assist him in engaging in safe falls prevention behaviour.

> “In both the acute and the rehab hospital, you know they have at the end of the bed… a whiteboard which says how much assistance you need for things. And they have their own little abbreviations, which the patient never understands. I had to explain to my dad what it meant!” (P39, carer).

#### Sub-theme. Family exclusion from conversations

Family members and older consumers were strongly critical of family being excluded from conversations about care. One older person stated “I would like it if I can talk to my family and they know what was going on. You don’t have to keep the family in the dark” (P23).

Carers stated that they needed to be included in education and communication. “You have to educate the carers. Absolutely. Family know their parents best, doctors and nurses need to listen and consider their viewpoints and not just go by the medical point of view” (P39, carer).

### Theme 5. Underlying attitudes of ageism

Some participants expressed feelings that their hospital care, including the inadequate communication about falls education received from staff was influenced by ageist attitudes. They felt that rather than being provided with tailored care, “…they lump you all together in the old person category” (P41). Subsequently, negative assumptions based on age appeared to question consumers’ capability and motivation to engage in their own care. “Older people would understand more if they weren’t treated like idiots. People treat you like you don’t know what’s going on, like you have dementia. They don’t seem to want to deal with the elderly” (P11). One focus group participant stated vehemently, “They’re not interested once you’re over that age group, it’s not fair” (P3). Family members also described engaging with staff who they felt displayed ageist attitudes, with one re-telling her mother’s experience. “…it was just some of the ways they spoke to her. They just treated her, you know, like she left her brain at the front door…and she was a registered nurse as well!” (P1, carer).

## Discussion

Older consumers’ feedback indicated they were challenged by sporadic and inconsistent falls prevention education and inadequate communication or support while in hospital. The inability to mitigate their risk of falling resulted in consumers feeling anxious and uncertain about how to undertake tasks, such as toileting safely, and they felt deterred from seeking information and assistance. Overall, older consumers’ feedback identified that communication and education about falls prevention was inadequate, and not what they expected. Participants who had fallen or nearly fallen, as well as those who did not fall while in hospital, reported similar experiences. These findings compare to a recent study conducted in the UK that found older adults who fell in hospital experienced inadequate communication, ageism and anxiety about maintaining their dignity, particularly when needing to use the toilet [41]. An Australian study also found that hospitalised older adults had limited knowledge about hospital falls risk and their experiences of falls prevention education were inconsistent [26]. Inadequate communication and education inadvertently resulted in consumers in the current study being motivated to maintain their independence even when they felt uncertain or unsafe regarding their planned actions, so as not to bother staff. Low awareness of falls, unwillingness to seek help and the desire to maintain their independence whilst in hospital are known to lead to older patients taking risks that could lead to falls [15, 42].

When viewed through the lens of the COM-B framework of health behaviour change[35], consumers’ reflections suggest that their hospital admission did not engender sufficient levels of capability, motivation and social opportunity to promote engagement in falls preventive behaviour [34, 35]. Health education that raises patients’ awareness of falls and develops positive motivation and opportunities to seek assistance is essential to improve older patients’ safety. Consumers in our study repeatedly stated that no one had raised their awareness that falls were a problem in hospitals and misperception of personal falls risk in hospitals has been confirmed as a significant problem [43]. As adult learners, many older consumers had useful prior learning and insight into falls prevention. Concepts of adult learning support using prior knowledge to engage learning and raise motivation about falls prevention while in hospital [44]. However, consumers reported that only a few staff explained the required strategies or listened to the older person’s input. A recent scoping review found that the design and delivery of falls education needs to be tailored to the individual’s needs, use a combination of education modes, and incorporate theories of health behaviour change and educational principles, including active learning, to better engage patients [17]. Falls guidelines indicate that education is an effective intervention rather than restrictive interventions such as alarms, [11] and integrative reviews on older patient’s safety and function conclude that maintaining and restoring function needs to be prioritised for older patients rather than encouraging restrictive strategies [45, 46].

Many consumers in our study reported that staff appeared too busy to promptly attend to patients’ care due to time pressure and competing demands. Framed within concepts of behaviour change theory, patients were confused by this apparent lack of opportunity (which health behaviour theory explains as both social and environmental) [34, 35] when they were struggling to mobilise, leading to distressing experiences of incontinence. These findings are supported by a review reporting that older patients value their dignity, prioritise their care over falls prevention and experience feelings of disempowerment, loss of independence and threats to perceived self-identity while in hospital [47]. Interactions by some staff reinforced feelings of anxiety and uncertainty, with some participants feeling upset or disempowered and deterred from seeking assistance, suggesting that hospital systems need to provide better tools for staff to deliver patient education [48]. Staff need to ask open-ended questions to understand patients’ thoughts and feelings about their recovery and mobility, which is important in encouraging help-seeking and motivation to undertake safe behaviours to avoid falls [15]. Some consumers perceived that ageism contributed to poor communication, which further undermined their sense of dignity, respect, and trust in the healthcare system, indicating that staff need to develop a holistic understanding about ageing that encourages them to treat older people with dignity [47].

### Strengths and limitations of the study

Data were collected and analysed using a methodology that included strategies to ensure rigour and trustworthiness [49]. This included method triangulation of data analysis by three researchers. Older patients are known to be reluctant to complain or speak up when provided with poor medical care [50]. In order to support older consumers to feel they could be honest and open, interviews and focus groups were led by independent researchers after participants were discharged from hospitals. Participants may also have felt more confident to express their opinions without fear of repercussion since they were not in a healthcare relationship with the researchers [28]. Participants included a heterogeneous sample of older adults with diverse medical and social characteristics and their carers, who provided an observer perspective and enabled data triangulation and rigour. Results from our study are from one health setting in Australia, although participants had been admitted to various hospitals. The consolidated criteria for reporting qualitative research (COREQ) guidelines were followed when designing, conducting and reporting the study findings to improve the quality and transparency of the reporting [51].

## Conclusion

Gaps in providing high-quality falls prevention education adversely affect older patients’ development of capability and motivation for falls prevention while in hospital, and lack of support reduces their ability to engage in safe behaviour confidently. Further research should focus on increasing collaborations with older consumers and seeking their active partnership in designing and implementing suitable fall prevention education as part of hospital safety programs. Such programs should focus on reducing ageism within the health system, evidence-based adult learning and health behaviour change frameworks. Hospitals ought to provide falls prevention education to older patients using personalised approaches that are tailored to the individual’s priorities and needs.

## Data Availability

All data produced in the present work are contained within the manuscript.

## Acknowledgements

AMH and SV conceived and designed the study with support from LF and MEM. SV and TW led the data collection with assistance from AMH. AMH led analysis and interpretation with support from SV, JFC, TW, and JC. AMH led the drafting of the manuscript with assistance from SV and JFC. All authors provided feedback on the manuscript drafts and read and approved the final manuscript submitted.

## Declaration

The authors have no conflicts to declare.

